# Statistical analysis of national & municipal corporation level database of COVID-19 cases In India

**DOI:** 10.1101/2020.07.18.20156794

**Authors:** Naman S. Bajaj, Sujit S. Pardeshi, Abhishek D. Patange, Disha Kotecha, K. K. Mate

**Affiliations:** Department of Mechanical Engineering, College of Engineering Pune, Savitribai Phule Pune University, Pune; Department of Electronics & Telecommunication Engineering, College of Engineering Pune, Savitribai Phule Pune University, Pune; Department of Mechanical Engineering, Pimpri Chinchwad College of Engineering & Research, Ravet, Pune, Savitribai Phule Pune University, Pune

**Author notes:** Corresponding Author: Abhishek D. Patange, Email ID.

**Keywords:** COVID-19, Pandemic, Statistical approach, Regression analysis

## Abstract

Since its origin in December 2019, Novel Coronavirus or COVID-19 has caused massive panic in the word by infecting millions of people with a varying fatality rate. The main objective of Governments worldwide is to control the extent of the outbreak until a vaccine or cure has been devised. Machine learning has been an efficient mechanism to train, map, analyze, and predict datasets. This paper aims to utilize regression, a supervised machine learning algorithm to assess time-series datasets of COVID-19 pandemic by performing comparative analysis on datasets of India and two Municipal Corporations of Maharashtra, namely, Mira-Bhayander and Akola. Current study is an attempt towards drawing attention to the dynamics and nature of the pandemic in a controlled locality such as Municipal Corporation; which differs from the exponential nature observed nationally. However, for limited area like the one considered the nature of curve is observed to be cubic for total cases and multi-peak Gaussian for active cases. In conclusion, Government should empower district/ corporations/local authorities to adopt their own methodology and decision-making policy to contain the pandemic at regional-level like the case study discussed herein.

## 1. Introduction

China Country office informed the WHO of unknown cases of pneumonia on 30^th^ December 2019. On investigation, the officers found a link to the Huanan Seafood market of Wuhan. Out of 44 cases, 33 patients were stable, but 11 were severely ill (WHO, 2020) [1]. Since its discovery, approximately 216 countries have been affected by it, with the total number of confirmed cases being 11.4 Million and 534K deaths. India currently has 697K cases and 19693 deaths, placing it at the third-highest position in the world as of 6 July (WHO, 2020) [2].

Coronavirus showed similar etiology to the Severe Acute Respiratory Syndrome (SARS) virus of 2006. Hence, it is also known as SARS-2 or COVID-19 (WHO, 2020) [3]. It is a contagious disease that spreads via minute respiratory droplets through coughing, sneezing, or close contact. Patients can experience the symptoms in 2 to 14 days of incubation, such as high fever, body pain or weakness, dry cough, breathlessness, pneumonia, kidney failure, and respiratory distress. In some cases, the patients do not show any symptoms of the virus; that is, they are asymptomatic. In both scenarios, testing of the patient is essential for verification, and they should be isolated (He et al., 2020; Singhal et al., 2020) [4] [5]. Thus to curb the rate of infection and prevent community spread wearing masks, to avoid crowded spaces and maintaining the protocols of social distancing is essential (WHO, 2020) [3].

In India, the first confirmed case of COVID-19 was of a man with travel history from Wuhan (WHO, 2020) [3]. Although the progression of cases was slow initially, the Indian government imposed a lockdown on 25^th^ March to prevent human-to-human transmission and contain the pace of the virus. After all the protective measures, the growth of the virus has continued at an exponential rate in the country Figure 1. Thus, modeling the current situation, analyzing the expansion pattern, identifying the reason for such behavior, and using it to forecast the next trend of the virus will help us develop coping mechanisms.

In Section 1, the current scenario has been explained along with the background and requirement of the proposed methodology. Section 2 details the importance of regression and the theory behind it. Details and motivation regarding the case study have been explained in Section 3. Section 4 consists of graphs and results after analysis. Section 5 details the comparative discussions of the obtained results followed by the conclusion in Section 6.

## 2. Literature review

Machine learning algorithms are used in all wakes of life, including the business sector, finance, medical, engineering, and educational domains. It is exceptionally efficient in the prediction of healthcare data (Mai et al., 2017, 2; Purcaro et al., 2018, 3; Ye et al., 2003) [6] [7] [8]. Right from helping understand the influenza pandemic dynamics to analyze data for prediction of the growth rate of COVID-19, its role is unmatched. As the cases of COVID-19 continue to at frightening rate, panic continues to spread among the people and their government. Ceased production, reduction in goods and services, and a collapsing economy and an increase in fatalities, create a situation of mass hysteria. In such scenarios, it can help countries make the right decisions and avoid loss of human lives and economy. Designing an accurate outbreak model will provide essential insights into new policy-making and will also help evaluate the result of the policies (Remuzzi et al., 2020) [9].

Several mathematical and statistical models have been utilized by researchers to study the behavior of the virus, people’s response, and predictions. Ramjeet, 2020 in his paper used six regression-based analysis models, i.e. quadratic, cubic, third, fourth, fifth, sixth degrees, and exponential polynomial on the dataset of India (Yadav, 2020) [10]. It provided a functional analysis of data, and the prediction model was viable for only seven days from the analysis. Employing Support Vector Regression method, Milind et al., 2020 carried out five different analyses, namely predicting spread, analyzing growth rate and types of mitigation, predicting recovery rate, the transmission of the virus, and correlation to weather conditions (Yadav et al., 2020) [11]. An ARIMA model with Exponential Smoothing and Holt-Winters model was used by Vikas et al., 2020 for regression analysis of India’s COVID-19 growth (Sharma et al., 2020) [12]. The model also underestimates the actual observations but can be updated to remove that issue, as suggested in the paper. It also predicted slowing in cases in the coming days, and proper guidelines and measures accompanied reduction in daily cases with it.

Building on the recent conceptualization of detecting connective communities in time series and Konstantinos et al., 2020 developed a novel spline regression model to determine knot using community detection in the complex network for Greece dataset (Demertzis et al., 2020, 5) [13]. Owing to diversity, the difference in geography, a large population, the study would not be directly beneficial in predicting growth rate for India. Poonam et al. used a linear and polynomial regression models on the datasets of various states of India (Chauhan et al., 2020) [14]. It highlighted a difference in national and state-level models, similar to the methodology proposed in this study. Gaurav et al., 2020 and Fairoza et al., 2020 have both performed analysis during the early onset of the corona (Gupta et al., 2020; BintiHamzah et al., 2020) [15] [16]. Gaurav, 2020 employed an SEIR and regression model with the data following a linear pattern. It also predicted that community spread would increase cases exponentially, which can be verified with the current models.

On the other hand, Fairuza et al., 2020, performed real-time data query and visualization using SEIR predictive modeling. Sina et al., 2020 did a comparative analysis of ML and soft computing models for predictive analysis on datasets from Italy, China, Iran, USA, and Germany (Ardabili et al., 2020) [17]. A comparison of various models revealed that MLP and ANFIS showed the best results and can be employed for long term predictions.

As can be seen from above, various types of methods have been used for analysis. However, it has been limited to a national approach with a limited study on state-level differences. Barely any form of study on district-level assessment is undertaken. As a difference in state and national level dynamics have been proved previously, this paper’s aim would be to explore Municipal-level differences, by mapping and evaluating, datasets of Mira-Bhayandar and Akola.

## 3. Regression scheme

Regression is a category of machine learning statistics tool. It utilizes supervised ML that employs an algorithm to understand the mapping of output data to input. Its objective is to map the approximate function as accurately as possible and train datasets. Analyzing data, identifying patterns and making corrections with human help or correction is possible with help of machine learning. These tools and algorithms are used by regression for mapping and prediction. Regressions models are employed to study the relationship between single or multiple independent variable(s) (X) and dependent variable(Y) involving unknown parameter(s) (β) (Sarstedt et al., 2014, 3) [18].

The reason for such an analysis is to find *β* that best fits the model. The benefit of regression analysis is that it can compute the relative strength of independent variables effects. Additional analysis can also predict the value of y for a given x in the sample size n. It includes modeling and analysis of variables to determine the most fitting algorithm that can explain their correlation.

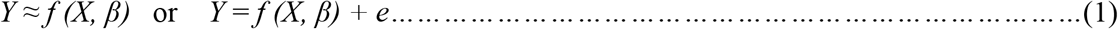

The term e represents error or residue. It is the distance between every observed value and its corresponding predicted value. ŷ(y-hat) represents prediction made by the best regression line. An assumption during this process is that e follows a normal distribution N (0, σ^2^) (Rasmussen et al., 2006, 1) [19]. Therefore, e is calculated by:

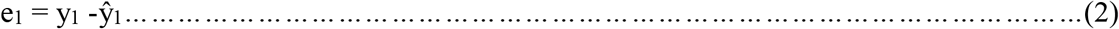

Calculating β uses the Least-Squares Linear regression method. It minimizes the value of e for the best fit curve. Another important quantity is the coefficient of determination or R^2^ that demonstrates the accuracy with which the proposed model describes the change in the dependent variable. It is represented by the formula:

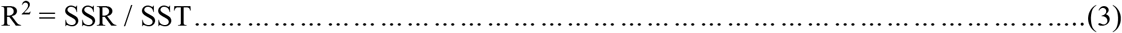

Where SSR represents square of difference between the regression line and average value’s sum (sum of the squares), and SST is the square of difference between each observation and the average value’s sum (total sum of squares),

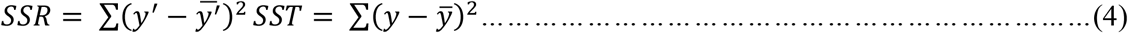

where Y_i_’s are the observed values corresponding to each X, the estimated (predicted) value of the response variable is represented by 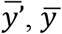 depicts the average of Y_i_, and n is the total observations. The value of R^2^ always lies between 0 and 1. For R^2^ = 1, the model has perfect determination and a value closer to 1 indicates a better model.

Fitting curves used for modeling are simple linear, multiple linear, polynomials of varying orders such as quadratic, cubic, exponential, Gaussian, and logarithmic. The analysis performed in this study employed cubic, exponential, and multi-peak Gaussian curves.

### 3.1 Cubic regression

The best fit line in this represents a curve rather than a straight line. It is much more flexible to incorporate data with complex relationships. It is useful when the plotted data first curves one way then another. It is considered a special case of the multiple linear regression models. The function that represents the graph is:

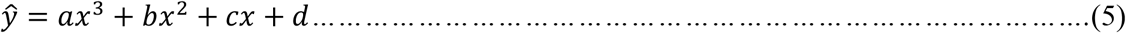

The following system of equations helps to find the value of coefficients:

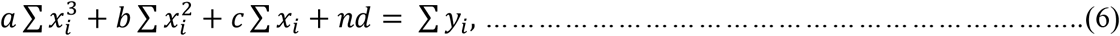

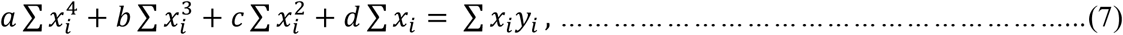

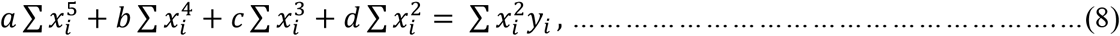

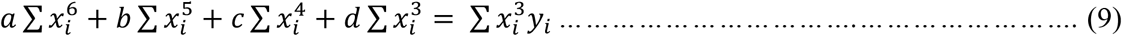

For comparing the efficiency, the model considers the value of standard error of regression (*Ā*), correlation coefficient (R), and coefficient of determination (R^2^).

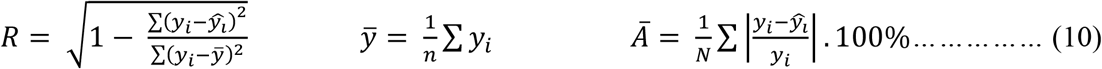

This method explains the district models in the study. It represented the total cases, the total number of deceased and total recoveries in the model.

### 3.2 Exponential regression

It is one of the simplest types of non-linear regression. When the coefficients vary non-linearly, this method gives the best fitting curve.

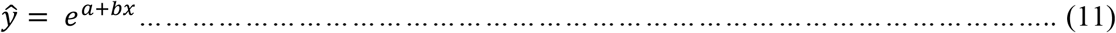

Coefficients ‘a’ and ‘b’ can be calculated with the help of:

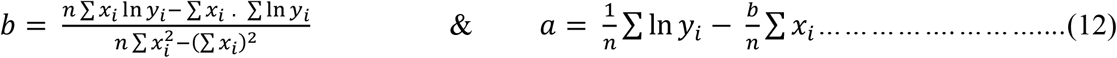

For the study, the country’s statistics in terms of total cases, recoveries, and deaths follow an exponential trend.

Similar to cubic regression, standard error of regression (*Ā*), correlation coefficient (R), and coefficient of determination (R^2^) help to understand the efficiency of the model used with the help of the same formula. If the correlation coefficient is 0, there is no relation between X and Y, for negative value they are inversely proportional, and they are strongly related for correlation coefficient’s value as 1. Both methods follow a similar path while solving the problem. It compares unknown data in a tabular format to empirical formulae and finds the closest match. It utilizes the Least Square Method to obtain the best fit for which residual is minimum.

### 3.3 Multi-peak fitting Gaussian

This method is especially useful as it increases the fit by the mathematical algorithm and numerical calculation to find Gaussian sub-peaks in the model. The sub-peaks are then combined, which results in a waveform that consists of multiple peaks (Qing et al., 2013) [20]. When the existing methods are unfit to describe the data or while the process involves small datasets, it can provide uncertain measures on predictions.

In this method of approach, first, a Gaussian process prior is assumed:

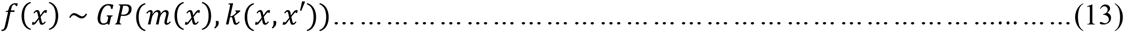

Here, k (x, x’) represents covariance function and m(x) is the mean function. A functional form does not limit Gaussian process regression (GPR) (Rasmussen et al., 2006, 1) [19]. Therefore, it calculates a probability distribution over all logical functions that can fit the data. For general purpose m(x) is taken as 0, since GPR is flexible to model mean arbitrarily well. One of the popular kernels used is a constant kernel composition with a radial basis function kernel. It encodes the similarity of inputs in space that corresponds to the similarity of outputs. It has two hyper-parameters, signal variance (σ²) length scale (l).

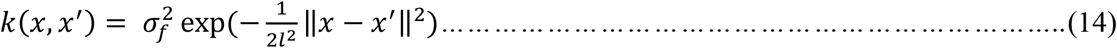

## 4. Methodology

The epidemiological statistics of India was obtained from the official COVID-19 website of India (Coronavirus India) [21]. The Indian subcontinent lies in the South Asian continent, with a population close to 1.380 billion. Maharashtra has been one of the most affected states during the pandemic, with approximately 217K confirmed cases as of 8 July. A regional level analysis and prediction of the outbreak would aid the officials in decision-making. The official websites of Mira-Bhayander and Akola corporations provided information regarding Coronavirus instances in the area, respectively (Mira-Bhayander; District Akola) [22] [23]. While Kerala’s official website was referred for details concerning the state figures for active cases and the number of people in quarantine from 30 January to 4 June (Kerala) [24]. Total cases, recovery, death, and active cases corresponding to 29 January to 25 May were examined for the national-level analysis. It is used as a benchmark for comparative analysis with other results and understanding. Akola’s statistics between 7 April and 5 June were computed, while those of Mira-Bhayander were between 27 March and 2 June. It comprised total positive cases in the regions and total cases, death, recovery, and number of cases daily.

SciDAVis (Scientific Data Analysis and Visualization), a cross-platform program for graphical presentation of datasets and data analysis, was used during the study’s analysis phase. It supports linear, non-linear and multi-peak functions and has built-in operations for column/ row statistics, convolutions, and filtering operations allowing with a user-friendly interface. Since the data needed non-linear analysis such as exponential, cubic and multi-peak Gaussian, this software was utilized for analysis.

## 5. Result of Regression Analysis

Results of analysis performed on SciDAVis using time series datasets of India, Kerala and Municipal Corporations of Mira-Bhayander and Akola have been stated in the form of graphical and tabulated format in this section. It forms the basis of the discussion done in Section 6.

### 5.1 Country-wise Plot

Fig. 1 shows a graph of total cases, recoveries, active cases and death for country wise dataset in the time frame 29^th^ to 25^th^ March. All four dotted lines mark the observed data while blue line represents the fit. It is an exponential fit obtained during regression analysis and can be represented with the equation: Y = Y_0_ + A * exp(x/t).

**Figure 1:**
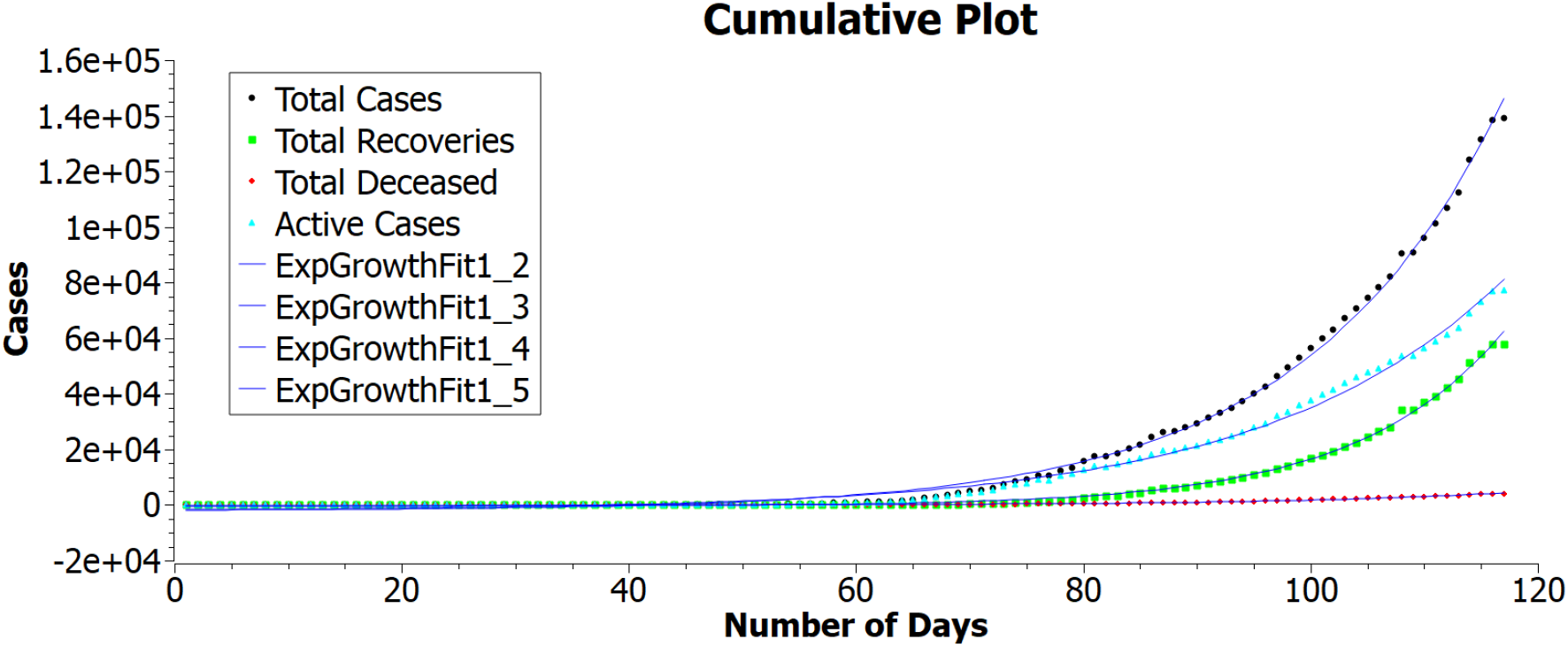
Cases in India from 29^th^ January to 25^th^ May

The R^2^ represents accuracy of fit, mentioned in Table 1, along with offset, amplitude and lifetime values. Mathematical values satisfying its equations have been mentioned Table 1. All the curves have more than 99% accuracy of fit.

**Table 1:**
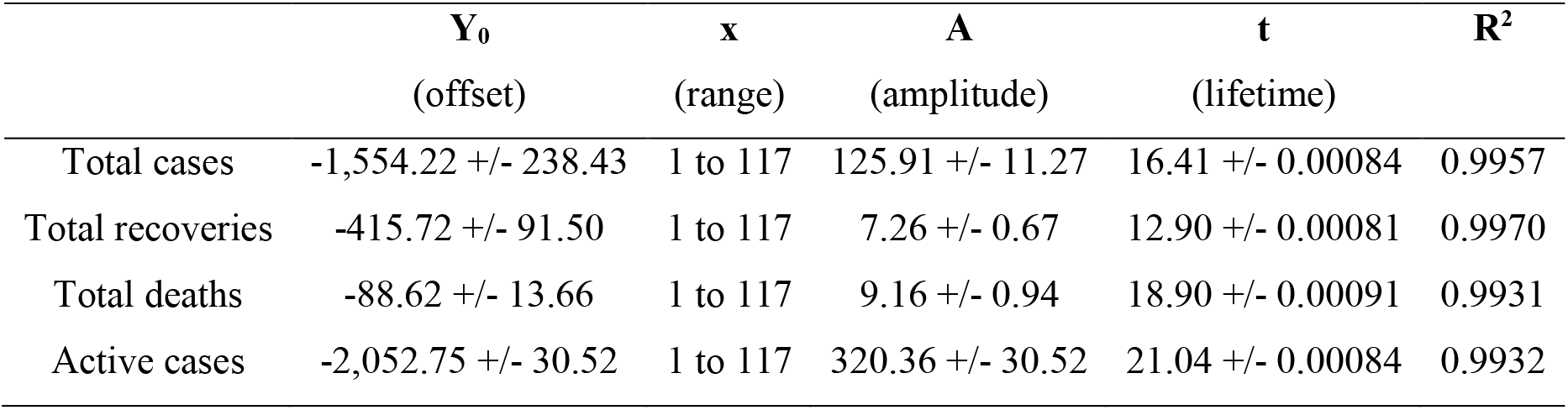
Values of offset, range of x, amplitude, lifetime and accuracy fit

**Figure 2:**
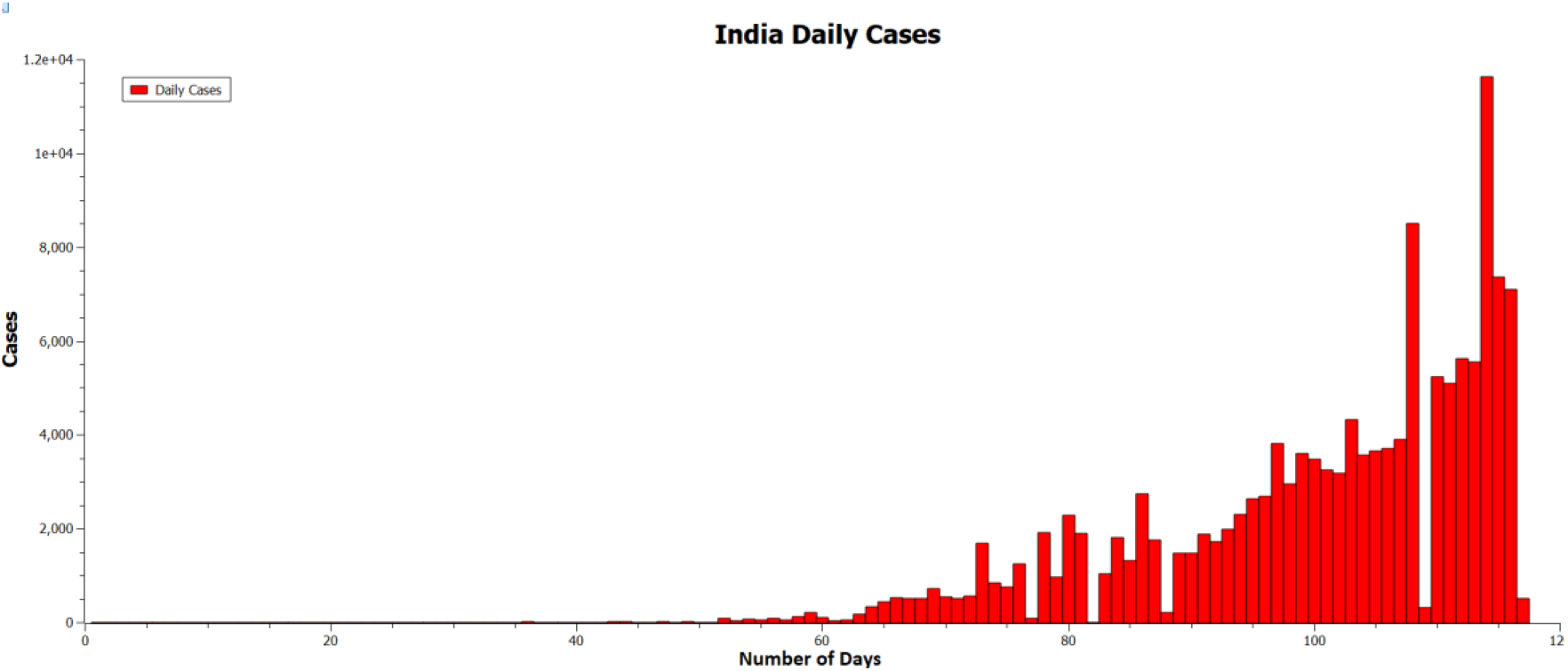
Daily positive cases in India from 29^th^ January to 25^th^ May

Figure 2 is a bar plot of daily number of positive cases in India between 29^th^ January and 25^th^ May. It has an exponential growth trend.

### 5.2 Region Plot

Figure 3 represents the cumulative plots of total deceased, cases and discharged for Mira-Bhayander between 27^th^ March and 2^nd^ June whereas Figure 4 represents the cumulative graph for Akola for the timeframe 7^th^ April to 5^th^ June. Dotted lines are the observed dataset for both the graphs while blue line is for cubic fit received. Unlike the exponential trend observed in Figure 1, both regional plots follow cubic regression curve. These curves can be represented by the equation: Y = a_0_ + a_1_*x + a_2_*x^2^ + a_3_*x^3^.

**Figure 3:**
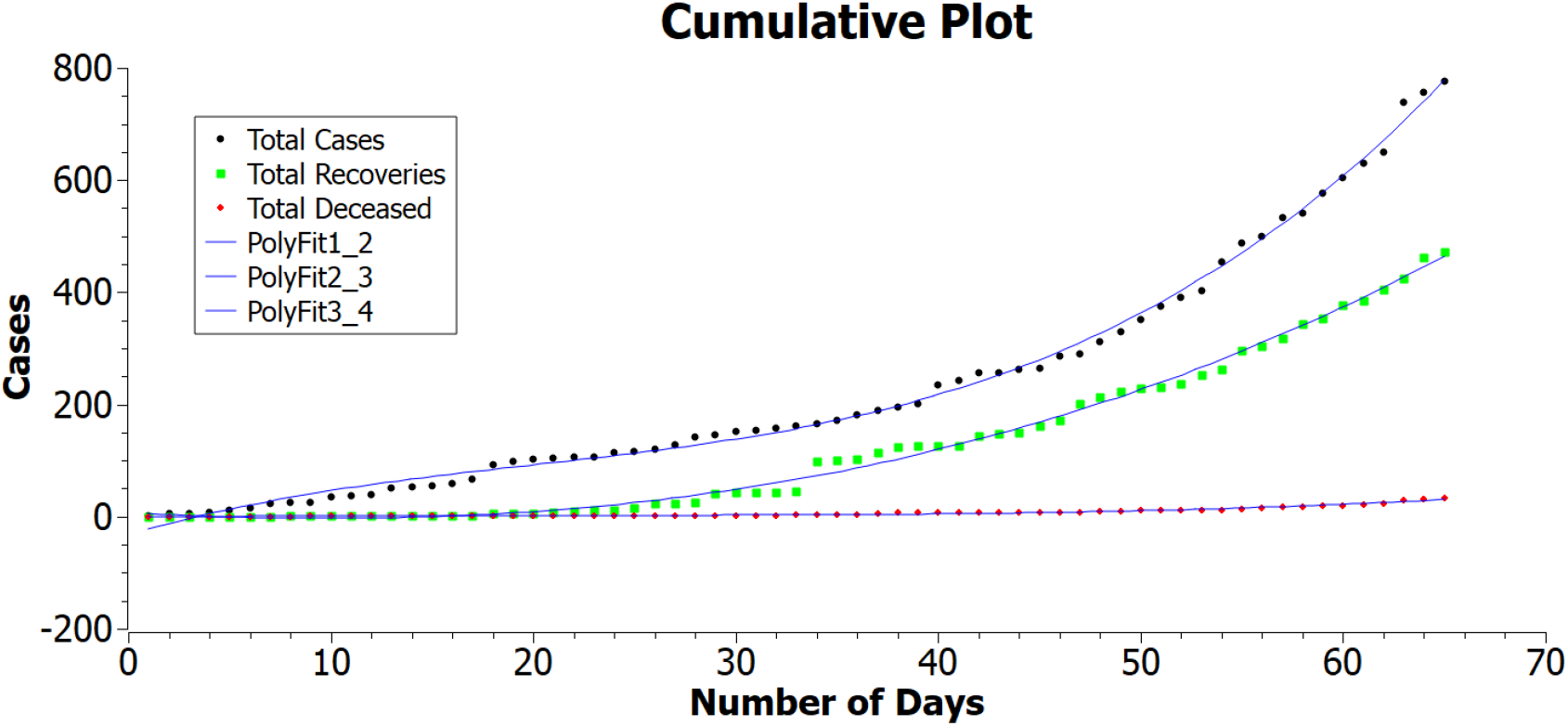
Cases in Mira-Bhayander from 27^th^ March to 2^nd^ June

**Figure 4:**
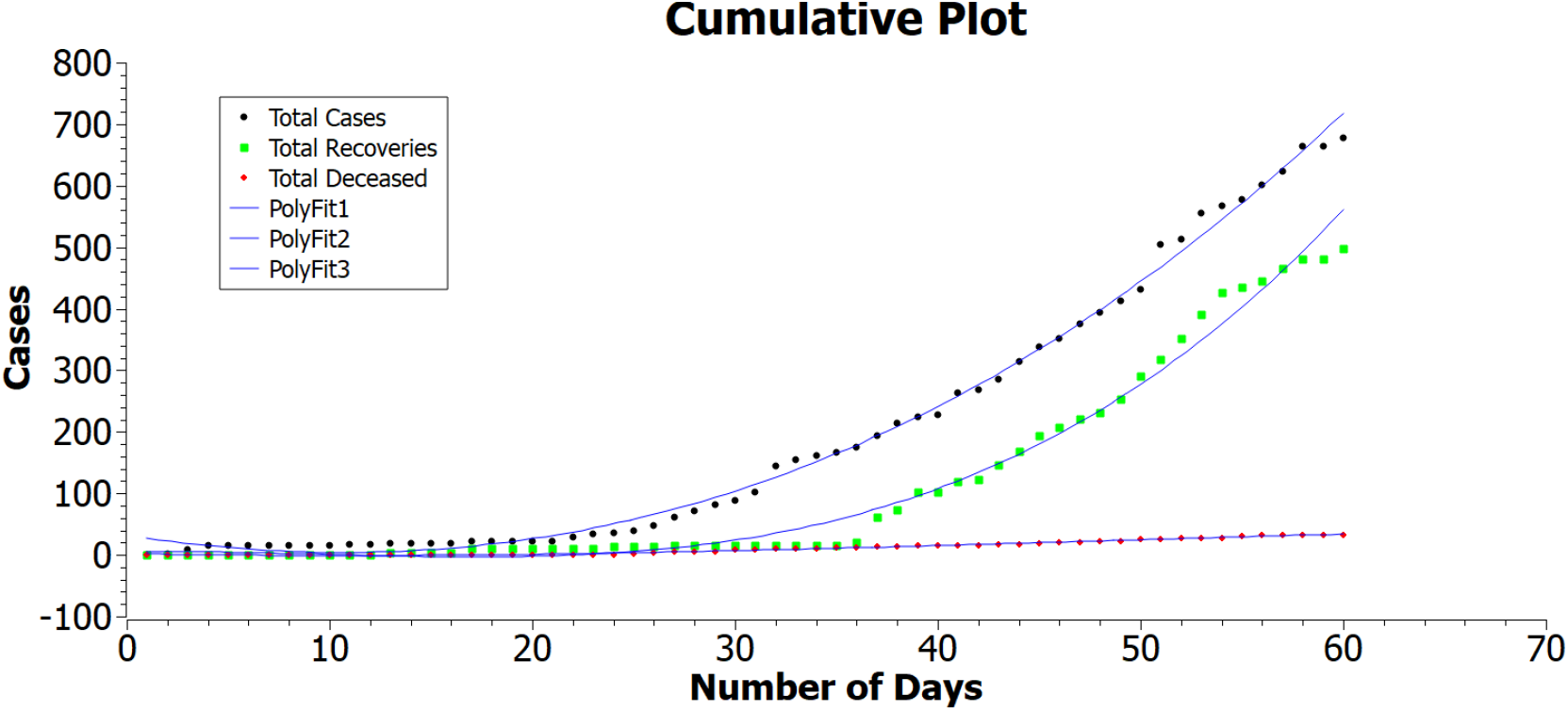
Cases in Akola from 7^th^ April to 5^th^ June

Accuracy fit (R^2^) for each curve in Figure 3 and 4 values have been tabulated in Table 2 along with values of coefficients.

**Table 2:** Values of coefficient and accuracy fit for Mira-Bhayandar and Akola

| Mira-Bhayandar | $a_0$ | $a_1$ | $a_2$ | $a_3$ | $R^2$ |
| --- | --- | --- | --- | --- | --- |
| Total cases | -32.80 +/- 6.50 | 10.65 +/- 0.84 | -0.33 +/- 0.029 | 0.0055 +/- 0.00029 | 0.9966 |
| Total recoveries | 7.10 +/- 5.20 | -2.01 +/- 0.67 | 0.09 +/- 0.023 | 0.00074 +/- 0.00023 | 0.9952 |
| Total deaths | -1.65 +/- 0.59 | 0.49 +/- 0.077 | -0.021 +/- 0.0027 | 0.00033 +/- 2.70 | 0.9810 |
| Akola | $a_0$ | $a_1$ | $a_2$ | $a_3$ | $R^2$ |
| Total cases | 31.68 +/- 7.95 | -5.096 +/- 1.11 | 0.22 +/- 0.042 | 0.00085 +/- 0.00045 | 0.9957 |
| Total recoveries | 4.59 +/- 10.79 | 0.50 +/- 1.52 | -0.13 +/- 0.057 | 0.0047 +/- 0.00062 | 0.9859 |
| Total deaths | 2.11 +/- 0.55 | -0.62 +/- 0.078 | 0.032 +/- 0.0029 | -0.00023 +/- 3.22 | 0.9924 |

Active cases datasets of Mira-Bhayandar and Akola is depicted in Figures 5 and 6, respectively. Both the graphs follow a 3-peak Gaussian curve unlike the plot of active cases of India in Figure 1 that resembles an exponential plot.

**Figure 5:**
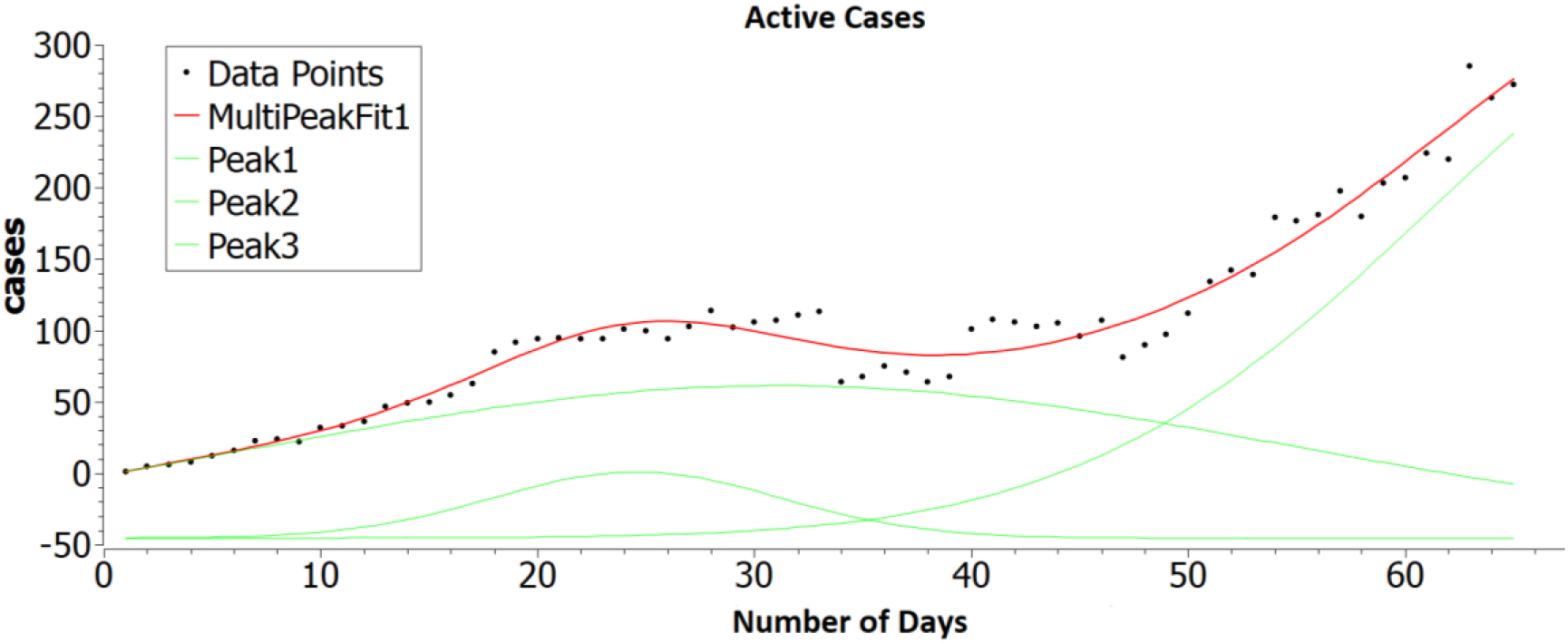
Active cases in Mira-Bhayander from 27^th^ March to 2^nd^ June

**Figure 6:**
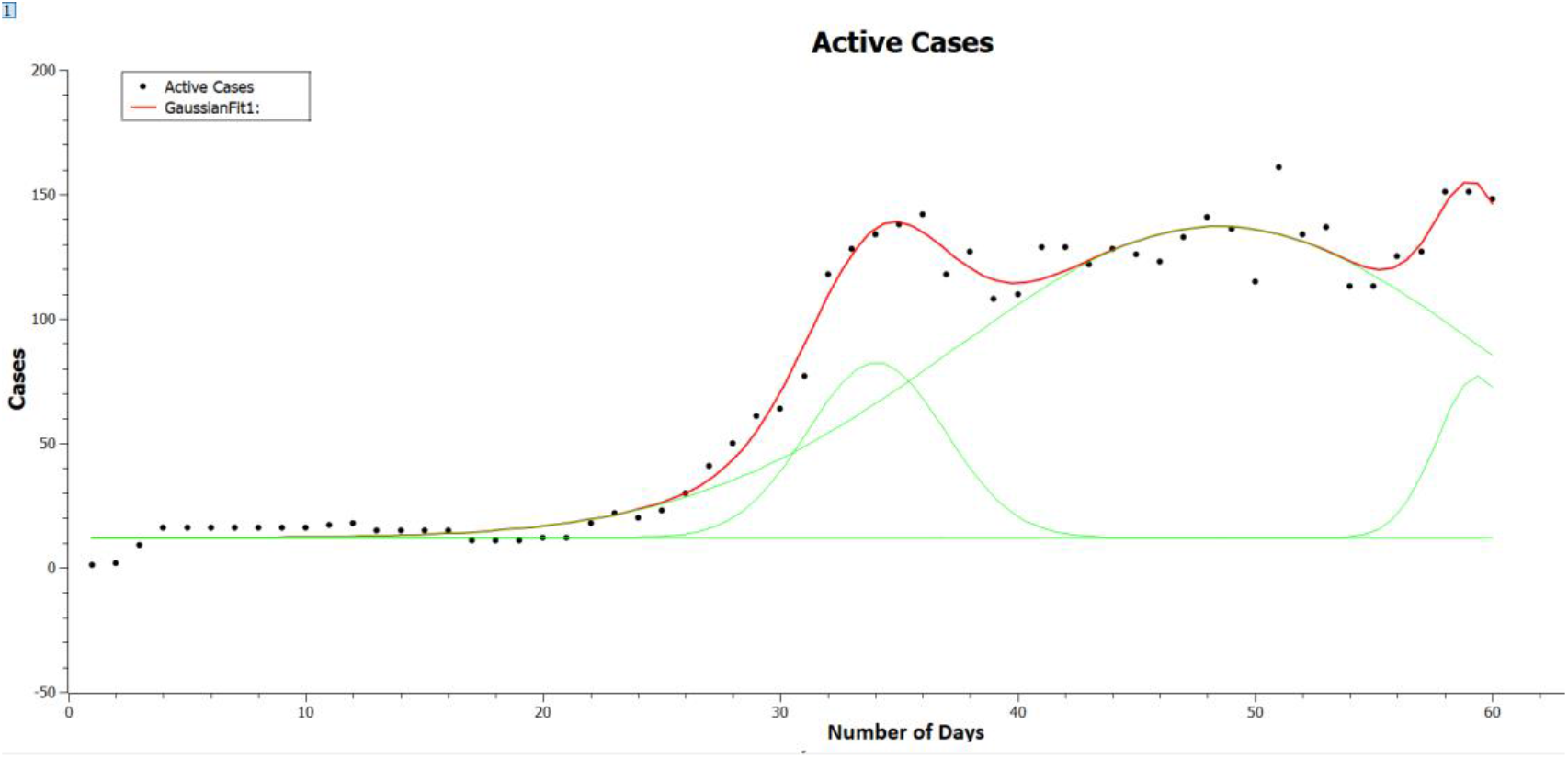
Active cases in Akola from 7^th^ April to 5^th^ June

Table 3 contains the tabulated data characteristics of each peak in Figures 5 and 6 that is values of area, centre, width, height and total accuracy of the fit.

**Table 3:** Values of area, centre, width, height and accuracy fit for Mira-Bhayandar & Akola

| Mira- Bhayander | Area | Centre | Width | Height | R<sup>2</sup> |
| --- | --- | --- | --- | --- | --- |
| Peak 1 | -4,897.11 | -14.48 | 41.67 | -93.74 | 0.9664 |
| Peak 2 | 820.94 | 24.65 | 13.41 | 48.83 |  |
| Peak 3 | 9,380.82 | 75.40 | 27.87 | 268.55 |  |
| Akola |  |  |  |  |  |
| Peak 1 | 513.098 | 34.04 | 5.81 | 70.44 | 0.9833 |
| Peak 2 | 3,497.83 | 48.48 | 22.30 | 125.13 |  |
| Peak 3 | 278.72 | 59.38 | 3.42 | 64.90 |  |

Figures 7 and 8 represent bar plot of the daily positive in the regions of Mira-Bhayander and Akola, respectively. Both figures record four significant spikes for particular dates. These spikes correspond to important lockdown dates.

**Figure 7:**
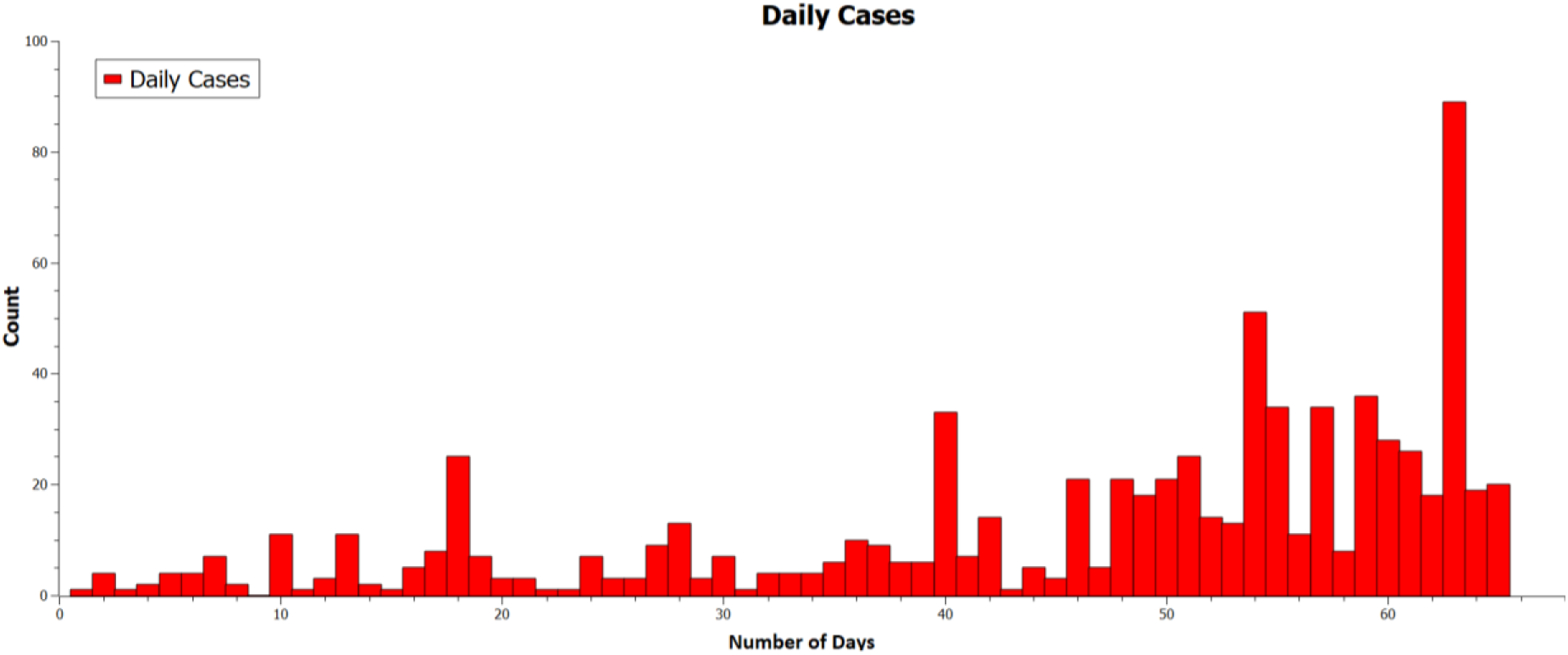
Daily positive cases in Mira-Bhayander from 27^th^ March to 2^nd^ June

**Figure 8:**
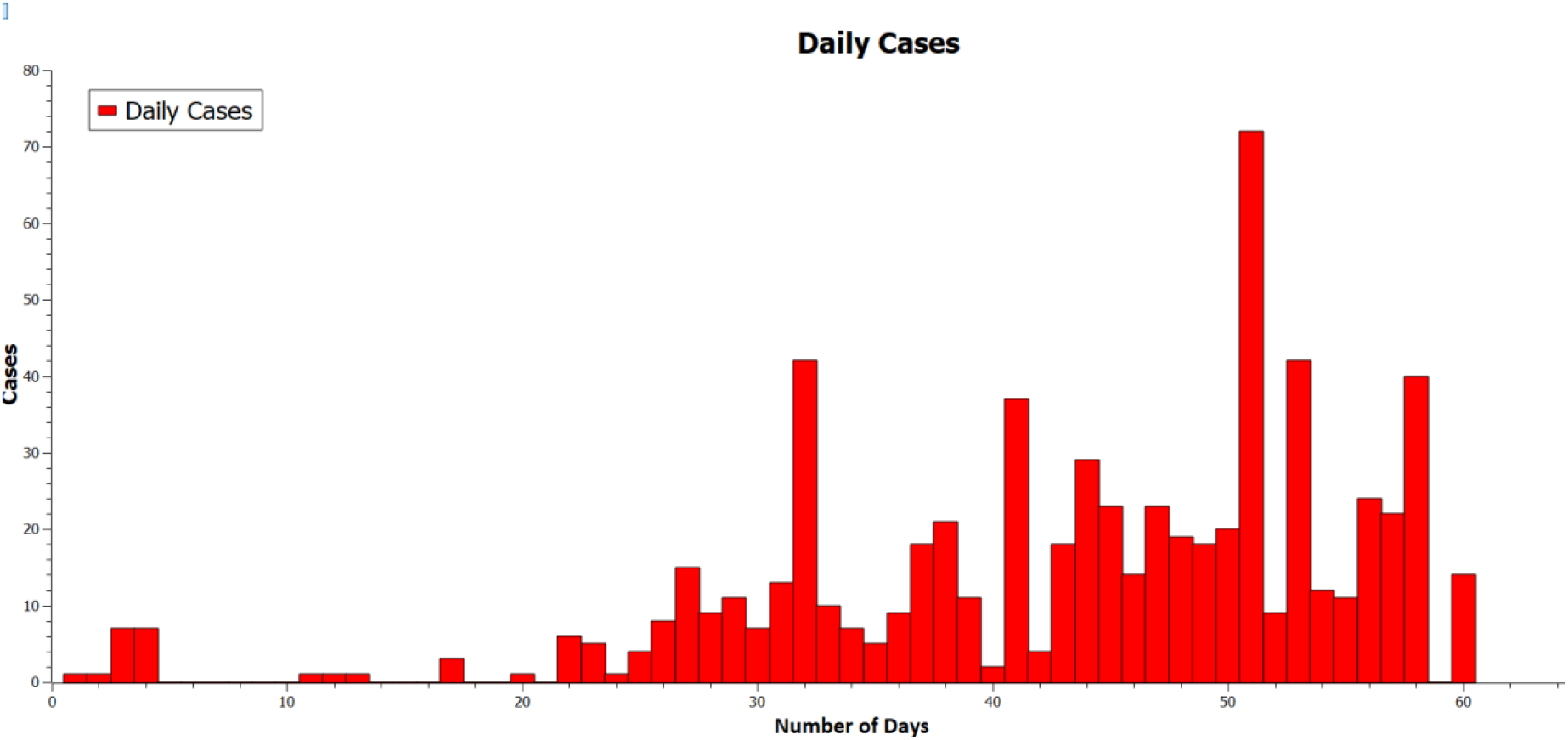
Daily positive cases in Akola from 7^th^ April to 5^th^ June

A comparative discussion on the significance of these spikes has been performed in the Section 5. Lockdown that was implemented earlier is a form of quarantine and since we had the data for number of people in quarantine in Kerala, this statistics was considered to compare with the case study of Mira-Bhayander and Akola. Figure 9 is the plot for active cases in Kerala between 30^th^ January and 4^th^ June. Total people on quarantine on each date in the state are mentioned in Figure 10.

**Figure 9:**
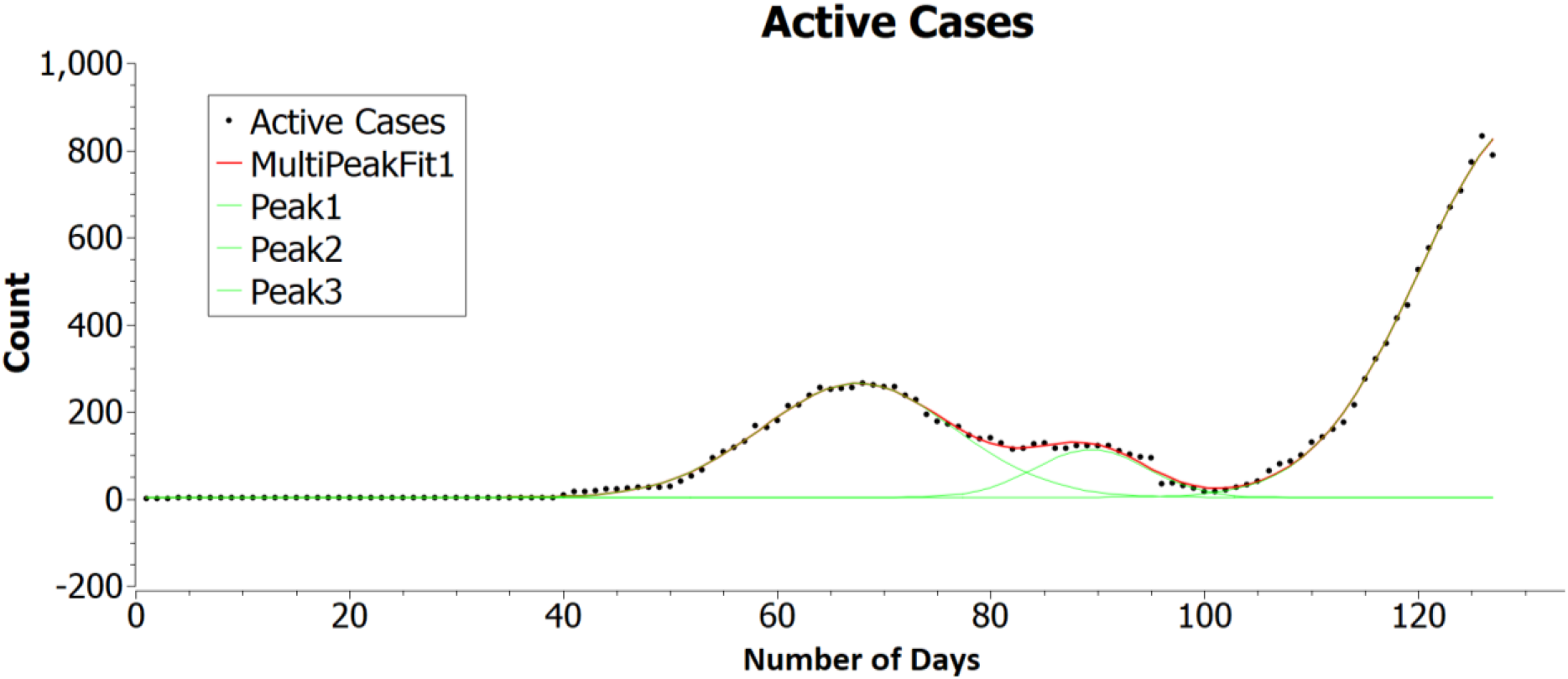
Active cases in Kerala from 30^th^ January to 4^th^ June

**Figure 10:**
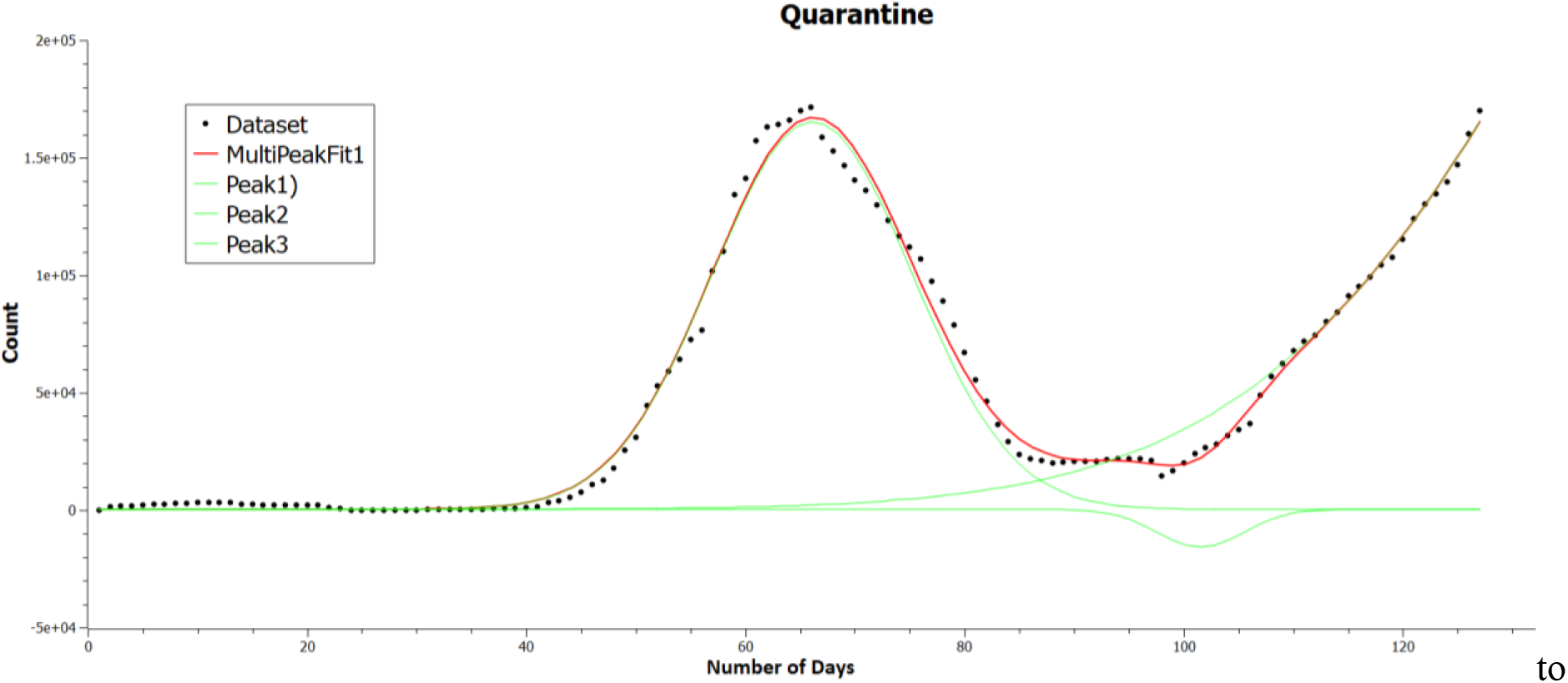
Quarantine cases in Kerala from 30^th^ January 4^th^ June

Plot of total active cases and number of people in quarantine follow a 3-peak Gaussian curve similar to regional active cases. Kerala was one the earliest state in India to be affected by the pandemic; the whole state underwent quarantine implementation. The plots obtained after analysis are also similar to regional curves rather than country plot. Hence, its results can be easily compared with the controlled containment zones. Values of area, centre, width and height each peak observed in both the figures along with their overall accuracy of fit has been tabulated in Table 4.

**Table 4:** Values of area, centre, width, height and accuracy fit for Kerala

| Active cases | Area | Centre | Width | Height | R<sup>2</sup> |
| --- | --- | --- | --- | --- | --- |
| Peak 1 | 5,903.62 | 67.57 | 18.022 | 261.36 | 0.9974 |
| Peak 2 | 1,443.92 | 89.35 | 10.50 | 109.64 |  |
| Peak 3 | 21,357.71 | 130.062 | 19.76 | 862.20 |  |

| Quarantine | Area | Centre | Width | Height | R<sup>2</sup> |
| --- | --- | --- | --- | --- | --- |
| Peak 1 | 3,774,787.57 | 66.11 | 18.24 | 165,060.71 | 0.9925 |
| Peak 2 | -160,946.98 | 101.52 | 7.95 | -16,134.18 |  |
| Peak 3 | 44,805,334.72 | 178.094 | 66.46 | 537,834.040 |  |

## 6. Inferences drawn and discussion

### 6.1 Country and Region Cumulative Plots

Comparison of Figure 1 of the cumulative country plot with the plots of Mira-Bhayander and Akola from Figures 3 and 4 shows the difference in the curve obtained i.e. while the country plot follows an exponential path, regional curve behaves in a cubic manner. It can be attributed to the reason that the outbreak’s statistics behave differently at a lower level than at the nation-level.

When all such statistics are combined, the resultant curve should resemble an exponential pattern. Thus, the dynamics of COVID-19 at a regional-level change drastically and should be noted while implementing policies.

### 6.2 Daily Cases

The daily onset of cases in the regions of Mira-Bhayandar and Akola from Figures 7 and 8 show four notable spikes in the bar plot outbreak. The dates of these spike, lockdown and unlock dates for comparison have been mentioned in Table 5. Table 5 exhibits that the spikes are either an immediate result of or due to the anticipation of lockdown/ unlock. This feature can only be seen in a regional plot, unlike the nation plot. Thus, it can be taken as a reference for future policies.

**Table 5:** Date of occurrence of spikes for (a) Mira-Bhayander; b) Relevant dates, Akola; c) Lockdown and unlock dates, India

| Mira-Bhayander | Akola | Lockdown |
| --- | --- | --- |
| 16 April | 7 May | 15 April |
| 7 May | 18 May | 4 May |
| 21 May | 28 May | 18 May |
| 31 May | 3 June | **Unlock:** 1 June |
| (a) | (b) | (c) |

### 6.3 Active Case Plots

Active cases plot in Figures 5 and 6 of Mira-Bhayandar and Akola, respectively, mimic a Gaussian curve with 3-peaks. It can be attributed to the fact that when the number of cases in a region increases, the number of people in quarantine increases proportionally. However, as the number of cases starts to fall, the quarantine number also decreases. As a result, more people get infected, giving rise to active cases.

A prime example of this feature is graphs of Kerala in Figures 9 and 10 depicting active and quarantine cases that follow this hypothesis. The validation of this response can be found by analyzing active and quarantine case plots of Kerala, which shows that both factors are co-dependent. Therefore, it is crucial to maintain high rates for quarantine, irrespective of the number of active cases, to avoid further outbreaks. Only when the total active cases start to dwindle quarantine regulations should be relaxed.

India had previously been making decisions at the national level. However, after the commencement of lockdown four, a state-based approach was followed where each state devised its policies and regulations. The state implemented zone based policies depending upon the extent of contamination. While drafting the paper Maharashtra State Government implemented another policy that empowers districts/ Municipal Corporation to take its decisions, showing a shift in trend from macro-level to micro-level policy making approach. It supports the proposed problem statement, and it can help them model their regional datasets for analysis. It can also be utilized to forecast further developments in response to decisions.

## 6. Conclusion & recommendations

In this paper, the nature of growth of COVID-19 total cases, deceased, recovered and actives cases with regional datasets from two Municipal Corporation was analyzed and the trends obtained are compared with the national one. Following conclusions are derived from the current study:

- From the analysis carried out different trends are obtained for the regional datasets than the national which is an indicator of pandemics distinct feature at local or zonal level.
- The trends for active cases plot is moreover driven by the policy matters such as changes in quarantine regulations, mapping of different zones within the locality, containment zones, implementation of lockdown etc.
- Spikes in daily cases observed in the regional plot are an attribute of human psychology combined with lockdown outcomes.
- This study suggests that more districts and states in every country should be given the authorization to make decisions regarding policies and lockdowns regulations depending on the outbreak’s nature in particular regions.
- Rules of quarantine should be adhered to strictly until the total active cases do not fall to a minimum value for a continuous period. It would also help reduce spikes in daily cases observed around significant days.
- It is a known fact that lockdown affect the nature of cumulated and active cases, however, only lockdown cannot be the only solution considering other factors of economy. Hence, it is recommended to further devise mechanisms locally to identify and distinguish micro-containment and volatile zones (hot-spots) using which administration can take curative actions on these zones identified.
- The authors’ advice that after locating the micro-containment zones, divide the locality into micro-containment zones and analyze and target local solutions.

## Data Availability

All datasets were acquired from official websites of respective location

https://www.covid19india.org/

https://www.mbmc.gov.in/master_c/important_information

https://dashboard.kerala.gov.in/

https://dio-akola.blogspot.com/

## Acknowledgements

The authors would like to thank the healthcare professionals, their allied teams and sanitation workers of COVID-19. Additional thanks to Municipal Corporations of Mira-Bhayander and Akola, Corporation Government of Kerala and India.

